# Awareness And Knowledge Of ‘The Kerala Healthcare Service Persons And Healthcare Service Institutions (Prevention Of Violence And Damage To Property) Amendment Act, 2023’: A Cross-Sectional Study Among Medical Students in South Kerala

**DOI:** 10.1101/2025.03.31.25324921

**Authors:** Anita Ann Thomas, Anna Mohanlal, Anu Elizabeth, A Harishankar, Gayathri Ponath Sukumaran, V R Reshma, Anjum John

## Abstract

**Introduction:** Increasing episodes of violence against healthcare workers (HCWs) are of growing global concern. The World Health Organization (WHO) reports 8% - 38% of HCWs having experienced physical violence at least once in their lifetime. Healthcare settings are recognized as high-risk workplaces by the WHO.

In response to widespread protests against the murder of a house surgeon at the Kottarakara Taluk Hospital in 2023, the government of Kerala strengthened the 2012 legislation with the Kerala Healthcare Service Persons, and Healthcare Service Institutions (Prevention of Violence and Damage to Property Amendment Act, 2023.

This study aimed to assess the awareness and knowledge about the Prevention of Violence and Damage Amendment Act, 2023 among medical students and evaluate the impact of an educational intervention on them.

**Methodology:** Baseline awareness and knowledge about the Act was assessed using a validated online questionnaire followed by an awareness session highlighting key provisions, legal implications, and protective measures under the Act. (Intervention). Post-class knowledge and awareness were reassessed.

**Results:** After the intervention, 87.3% of participants demonstrated good knowledge, 6.9% moderate and 5.7% poor knowledge of the Act.

**Conclusion:** The study highlights the effectiveness of awareness classes as educational interventions in improving medical students’ knowledge of legislation that directly impacts them. Such initiatives prepare them to address future workplace challenges and foster safer healthcare environments.

## INTRODUCTION

Healthcare workers (HCWs) play a pivotal role in the welfare and health of society. They provide day to day care, manage emergencies, and act as frontline workers safeguarding public health. Despite this, their professional lives are fraught with risks and challenges, often enduring verbal abuse and physical violence from patients, their attendants, or even colleagues, instead of the appreciation they deserve. Such intimidations not only jeopardize their physical and mental wellbeing but also hinder their ability to perform their duties without fear. Violence is not only a breach of their fundamental right to safety but also endangers patient care, creating a ripple effect across the healthcare system.(1,2) In view of increasing episodes of violence against healthcare workers (HCWs) worldwide, the World Health Organization (WHO) asserted that healthcare has become a high-risk workspace for HCWs all over the world. It is reported that around 8% - 38% of HCWs have experienced physical violence at least once in their careers. Healthcare Workers face not just physical abuse but also verbal assault from patients or their attendants. Doctors and nurses involved in patient care besides emergency room staff are most at risk. (3)

India has witnessed a surge in violent incidents against HCWs, ranging from verbal to physical assaults, particularly in critical care and emergency settings.(4)The COVID-19 pandemic has worsened the situation with frustration over healthcare outcomes and misinformation regarding COVID-19, fuelling aggression. (5) Violence against healthcare workers is driven by multiple systemic factors, including insufficient consultation time, long waiting hours, understaffing, overcrowding, and inadequate security measures, which contribute to patient frustration and aggression. Additionally, lack of trust in the healthcare system, unrealistic expectations of treatment outcomes, misunderstandings about care, dissatisfaction with management practices, and unprofessional behaviour by healthcare workers further exacerbate tensions. Mental health issues, heightened stress among patients and bystanders, as well as untreated substance abuse and intoxication, also play a significant role in escalating violence in healthcare settings.(6) Addressing these challenges needs a multi-prong strategy with systemic improvements to ensure safety and wellbeing of the medical community.

Following large-scale protests against the tragic and brutal murder of a young house surgeon at Kottarakara Taluk Hospital, a Public Interest Litigation (PIL) was filed in the Kerala High Court, urging the state government to safeguard the work environment for healthcare workers. As a culmination of a cascade of events that followed, an Amendment was introduced to the pre-existing **The Kerala Healthcare Service Persons And Healthcare Service Institutions (Prevention Of Violence And Damage To Property) Act, 2012** through an ordinance to strengthen protection for doctors and healthcare workers. The modified Act is one of the strongest State laws in India for safeguarding the rights of Health Care Workers. This amendment sought to strengthen penalties, streamline reporting mechanisms, and ensure accountability for violence against HCWs. (7) It defines violence as “activities causing any harm, injury, or endangerment to life, or intimidation, obstruction, or hindrance to any healthcare service person in the discharge of duty, in any healthcare service institution, or damage or loss of property in healthcare service institutions.” Under this Act, offences are categorized as cognizable or non-cognizable and bailable or non-bailable, depending on the nature and severity of the incident. (7) The Act recognises any form of violence against HCWs, as a violation of the basic human rights of an individual. It acknowledges the severe physical and psychological impact on victims of violence, which diminishes their job commitment and overall wellbeing. Healthcare service institutions face reputational and financial damage as a collateral negative repercussion of violent incidents. Every healthcare worker operates under immense pressure in high-stress environments to provide the best care to patients, making workplace violence an urgent concern.(7,8) The Kerala Amendment of the 2012 Act provides a ‘Code Grey’ protocol, to manage aggressive or violent behaviour from anyone, in healthcare institutions. This code alerts security personnel to take immediate action, against threatening or abusive conduct, ensuring a swift response to potential incidents, allowing for faster resolution of such incidents. Kerala is the first state in India to implement the Code Grey system, according to its Health Minister. (9) To ensure swift delivery of justice, the Ordinance mandates the appointment of an investigating officer (not below the rank of Inspector), who must complete the investigation of the violent act within 60 days, from the registration of the FIR. Additionally, trial proceedings should be completed within one year, although the court may extend this period by six months if necessary. Special courts and designated prosecutors are assigned in each district to expedite the legal process.

The gap between policies and practice needs to be bridged for effective enforcement. The existence and scope of the guidelines for protection must be brought to the notice of the public for better implementation. Low awareness about legislations that safeguard their rights can deter healthcare workers from reporting violence or seeking help from the legal system, leading to psychological stress. Mere enactment of laws does not serve any purpose; these laws must be communicated, understood, and actively followed by the community. A lack of awareness about laws protecting their rights may discourage healthcare workers from reporting violence, while the “perceived” absence of legal recourse can contribute to their psychological stress.(10) Raising awareness about these legal protections is essential for encouraging reporting, reducing violence, and ensuring compliance with the law.(11,12) Despite its noble intent, the awareness and understanding of this Act among medical students—the future HCWs—remain underexplored. Medical students, often exposed to high-stress environments during their training, are particularly vulnerable to workplace violence. Familiarizing them with the provisions of this Act is critical, as it empowers them to recognize and respond to violence, understand their legal rights, and advocate for systemic changes within healthcare institutions.

An extensive literature review revealed that though there are frequent media reports of violence against health care workers, scientific studies on violence against HCWS are limited, particularly on the legal aspects and protections against violence. Santro Vento et al. identified emergency departments, mental health units, drug/alcohol clinics, and ambulance services as high-risk settings for violence. They identified physical harm, post-traumatic stress disorders, depression, reduced work interest, and absenteeism as consequences. They recommended infrastructure improvement within healthcare facilities, better staffing, improved communication skills, and reporting mechanisms.(13) Chinese researchers, Shukun Yao et al., highlighted escalating violence against HCWs in China since 2014, citing instances of extreme violence like murder and assault. They proposed legal reforms, countering false media reports, and improving doctor-patient communications. (14)Neeraj Nagpal et al., attributed violence in Indian hospitals to systemic issues like poor healthcare infrastructure and lack of facilities, especially in underfunded government hospitals and small private clinics, which were both deemed vulnerable to violence.(15)A study by Khiyani et al. investigated the prevalence and nature of workplace violence against healthcare workers (HCWs) in two hospital settings in Sagar, India, and found that a significant proportion of HCWs experienced verbal and physical abuse, with higher incidents reported in the government hospital compared to the private hospital. The study underscored the need for effective strategies and policies to protect HCWs from workplace violence. (16)The 2017 position statement by the Academic College of Emergency Experts & Academy of Family Physicians of India reported rising incidents of assault on HCWs, particularly in Emergency Care. Recommendations included developing Emergency Medicine Departments as standalone departments, emergency training for all physicians, improved communication skills, and countering negative media portrayals. (17)

A case study by Dr. George Jacob described the 2023 murder of Dr. Vandana as a failure of the system to protect its protectors, warning that future incidents would occur unless systemic reforms were embraced. The victim of the 2023 murder in Kerala, was portrayed as a “sacrificial lamb at the altar of a failed system”. Kerala recorded 469 attacks on medical staff and 70 cases of assault on hospitals since 2012.(18)

While the Amended Act criminalizes violence against HCWs and property damage, gaps remain, including there being no time limit for FIR filing, lack of provisions in legislation for habitual offenders of violence, and no mental health assessment of perpetrators. (7) This study aims to assess the knowledge and awareness of medical students regarding the Kerala Healthcare Service Persons and Healthcare Service Institutions (Prevention of Violence and Damage to Property) Amendment Act, 2023 and identifies gaps in legal and ethical training. It underscores the need for targeted interventions, such as workshops, simulation exercises, and policy discussions, to promote legal literacy and a culture of safety in healthcare settings. By evaluating knowledge improvement, post intervention, the study aims to inform policy makers and educational institutions for HCWs protection and healthcare system strengthening.

## METHODOLOGY

### STUDY DESIGN

This research was an intervention study, designed to measure the baseline awareness and knowledge of medical students about the 2023 Act, followed by an educational intervention and subsequent evaluation to determine the improvement in their understanding.

### STUDY SETTING

The study was carried out at PIMSRC, a premier medical institution in South Kerala. The setting provided access to a diverse population of medical students undergoing rigorous academic and clinical training, making it ideal for evaluating awareness about the Act.

### STUDY DURATION

The study was conducted over a period of three months, starting from the date of approval by the Institutional Ethics Committee. This time allowed for sufficient planning, execution, and evaluation of the intervention, including pre- and post-tests.

### SAMPLE SIZE

Since, at the time of conception of this study, there were no similar studies published, the proportion of students aware of the Act was assumed to be 50% (the maximum proportion) and calculated the sample size using the formula:

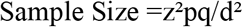

While keeping the relative precision to be 20% of the proportion, the sample size was calculated to be 196.

### STUDY POPULATION

All medical students willing to participate in the study, attend the awareness session, and complete the post test, were included in the study.

### SAMPLING PROCEDURE

The study population included all medical students enrolled in the medical college by universal sampling.

### METHOD OF DATA COLLECTION

A self-administered validated questionnaire was used to assess the baseline awareness and knowledge of participants regarding the Act. A consent form was attached to the questionnaire. Completion of the survey was taken as consent to participate in the study. The questionnaire consisted primarily of knowledge-based questions evaluating students’ understanding of key provisions, legal implications, and enforcement mechanisms of the Act. A subset of questions assessed awareness, including familiarity with the existence and intents of the legislation. After scoring their answers, a short educational session on the Act was held. This consisted of a structured awareness session covering the rationale, key provisions, legal protections, reporting mechanisms, enforcement strategies, and the role of healthcare professionals in mitigating violence in healthcare settings. After the intervention, a post-test was administered using the same questionnaire to evaluate changes in knowledge levels. To ensure comparability, the interval between the pre-test and post-test was standardized, with the post-test conducted immediately after the pre-test.

DATA COLLECTION TOOLS: A pretested and predesigned self-administered questionnaire was used for data collection. The questionnaire was scrutinised for validity and reliability (Cronbach’s Alpha = 7) after a pilot test. The revised questionnaire was printed and distributed among the participants along with consent forms.

### OUTCOME MEASUREMENT

The primary outcome of the study was the proportion of medical students demonstrating baseline awareness of the *Kerala Healthcare Service Persons and Healthcare Service Institutions (Prevention of Violence and Damage to Property) Amendment Act, 2023*. The secondary outcome assessed the percentage change in awareness levels among the study population post-intervention.

### DATA ANALYSIS

The responses were scored based on a pre-decided rubric. Total scores were calculated for each participant in both the pre- and post-tests. To find the knowledge level of each participant pre-test and post-test, mean scores of both the tests were taken as reference points for interpretation of the awareness levels as good and poor. Paired t-tests were conducted to assess whether the difference in mean scores was statistically significant. A p-value < 0.05 was considered to be statistically significant.

## RESULTS

In this interventional study, 245 participants attended both pre and post-tests. Since the sampling was universal and keeping the spirit of study in mind, we did not exclude the students who were willing to join.

### Demographic features of the population

The mean age of the participants was 21.07 years with a standard deviation of 1.777 years. Among the research population, 73.5% were female and 26.5% were male. Among the total of 245 participants, 2.4% belonged to the final year batch (batch of 2020), 37.1% belonged to the 2021 (pre-final) batch, 40.4% belong to the (first clinical) batch and 20% belong to the 2023(basic sciences) batch.

### Awareness

To assess awareness of participants about the existence of the Act, they were asked: “Have you heard about *The Healthcare Service Persons and Healthcare Service Institutions (Prevention of Violence and Damage to Property) Amendment Act, 2023*?” The results highlights that a majority (62.4%) of the study population were unaware of the Act, while 37.6% were aware of it.

### Knowledge questions

**Table 1:**
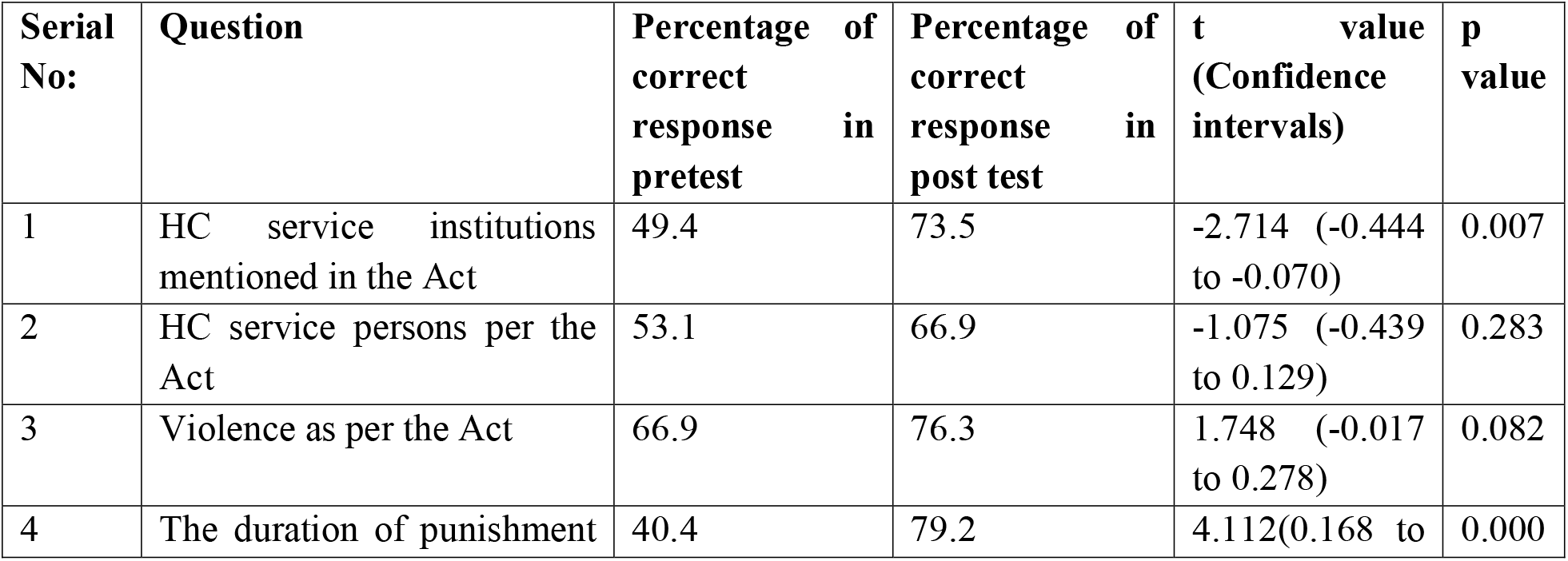
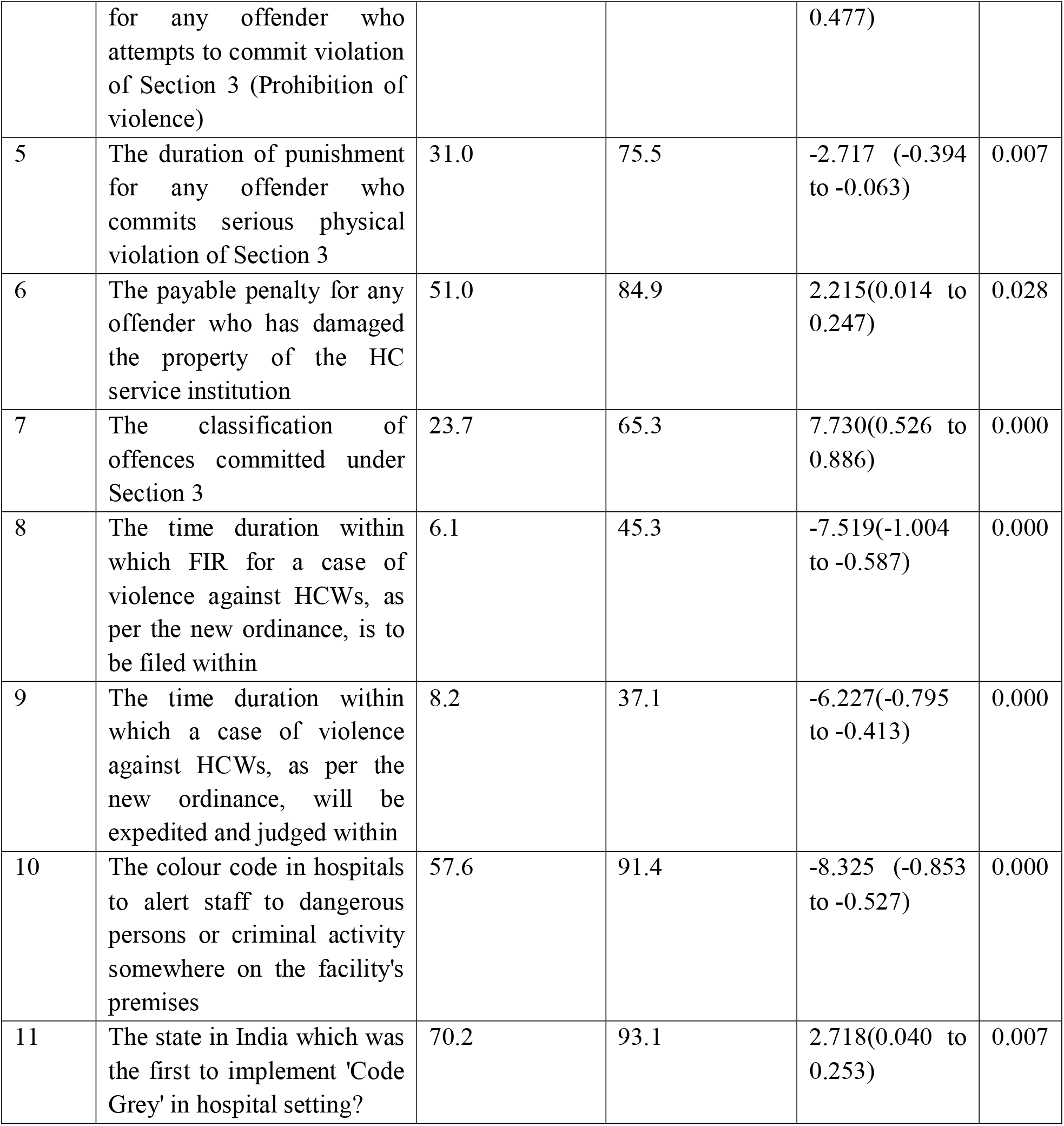
Table comparing the pre-test and post-test responses of questions related to Knowledge about the “Act”.

**Figure 1:**
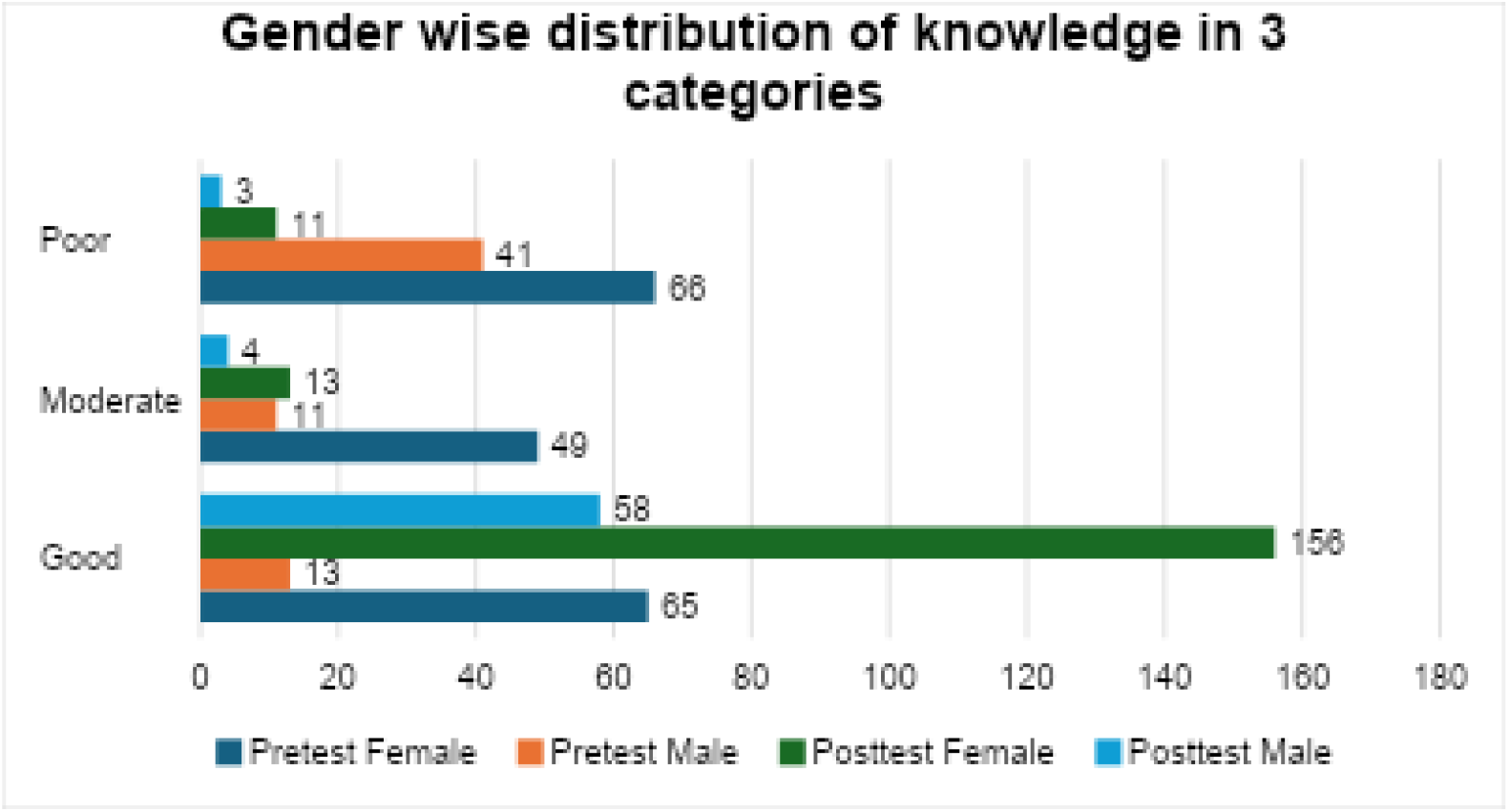
Showing the gender wise distribution of knowledge in 3 categories.

**Table 2:** Table of the gender wise distribution of pretest and post-test knowledge.

| Knowledge | Pretest Knowledge Female and Male | Post test Knowledge Female and Male |
| --- | --- | --- |
| **Good** | F 65 | F 156 |
|  | M 13 | M 58 |
|  | Total 78(31.8%) | Total 214 (87.3) |
| **Moderate** | F 49 | F 13 |
|  | M 11 | M 4 |
|  | Total 60 (24.5%) | Total 7(6.9%) |
| **Poor** | F 66 | F 11 |
|  | M 41 | M 3 |
|  | Total 107(43.67%) | Total 14(5.71%) |
| **Total** | F 180 | F 180 |
|  | M 65 | M 65 |
|  | Total 245 | Total 245 |

#### Attitude regarding training programs

**Table 3:** Table depicting distribution of study population based on their attitude towards mandatory training programs and awareness of their legal rights and protections.

|  |  |  |
| --- | --- | --- |
| Question 1: Should there be mandatory training programs for healthcare professionals to ensure a comprehensive understanding of the amended Act? |  |  |
|  | Pre – Test | Post-Test |
| Yes | 221(90.2%) | 232(94.7%) |
| No | 24(9.8%) | 13(5.3%) |
| Question 2: To what extent do you think healthcare professionals are generally aware of their legal rights and protections under the amended legislation? |  |  |
|  | Pre – Test | Post-Test |
| Fully Aware | 38(15.5%) | 53(21.6%) |
| Partially Aware | 158(64.5%) | 187(76.4%) |
| Unaware | 49(20%) | 5(2%) |

**Table 4:** Table showing Mean scores and Standard Deviation in Pre-test and Post-test knowledge Vs Age.

| Parameter | Pretest score out of 12 | Post tests score out of 12 |
| --- | --- | --- |
| Mean | 4.78 | 8.83 |
| Standard Deviation | 1.673 | 2.345 |

## DISCUSSION

### Key Findings and their Significance

This study assessed the awareness and knowledge of 245 students at a private medical college in Kerala, regarding “The Kerala Healthcare Service Persons and Healthcare Service Institutions (Prevention of Violence and Damage to Property) Amendment Act, 2023.” Prior to the intervention which included an awareness class after the pre-test, 37.6% of participants were aware of the Act, and 62.4% were unaware. The pre-test results revealed that 31.8% of the students had good knowledge, 24.5% had moderate knowledge, and 43.7% had poor knowledge, revealing a critical gap in legal knowledge. Following the educational intervention, the post-test results showed a significant improvement: 87.3% having good, 6.9% having moderate, and 5.7% having poor knowledge, respectively.

The mean pretest score was 4.82 increased to 8.83 with a statistically significant improvement with a statistically significant improvement (t = −22.834, p = 0.000), indicating that structured educational sessions can significantly enhance students’ understanding of protective healthcare laws like the Act.

This study reinforces the necessity of embedding legal awareness programs within medical education curricula, given the increasing incidence of violence against healthcare workers (HCWs). dditionally, there is a significant dearth of studies from Kerala or India that medical professionals’ knowledge of protective laws, making this study a valuable contribution to the existing literature.

### Insights from Existing Research

Workplace violence in healthcare is of well-documented global concern. According to the World Health Organization (WHO), 8%–38% of HCWs experience workplace violence, with underreporting being a major issue due to fear of retaliation and inadequate legal knowledge. (19) In India, studies indicate that over 75% of the doctors have faced at least some form of workplace violence, yet awareness of protective laws remains low. (20)A report by Jayadevan in 2019 highlighted the increasing frequency of violence against HCWs and the lack of strong legal enforcement, further underscoring the need for greater awareness and education on protective laws.(21) Attendants of patients committed 68.33% of the violence. Around 19 states in the country as of now have protection laws in place. (22,23)

A study by Divya Reddy et al. reported that only 166 (41%) of MBBS students were aware of the national and state laws protecting doctors, mirroring this study’s findings. In addition, 77.8% of students recognized that workplace violence includes physical assault, psychological violence, including verbal abuse, telephonic threats, and warning gestures, and vandalism, amounting to damage to medical equipment, and clinical establishments. The perceptions of students about violence widely varied, with one student (0.2%) perceiving violence as solely verbal abuse, five considering it a combination of verbal and physical abuse; twelve (3%) defining violence as only physical assault and seventy students (17.3%) viewing it as a varying mix of these elements. Surprisingly, two individuals (0.5%) did not associate any of these components with aggression toward doctors.(24)

According to a 2017 study by Ranjan et al., instances of vandalism have increased between 2014 and 2016. These acts of aggression have happened in private settings as well as government hospitals. About all the students (99.8%) knew that the number of violent crimes against doctors had increased. They learned about these incidents from the media in 91.6% of cases, acquaintances, and coworkers in 6.7%, and relatives in 0.5% of cases. Even while the role of the media was crucial in raising awareness of these incidents, it has been notorious for distorting the news for sensationalism. Just a very small fraction of 1.2% of the participants learned about this problem from their college instructors or professors. (25)

Another study by Krishnan et al. among dental surgeons found that 30.4% had experienced violence at their clinic, with 39.3% reporting verbal abuse, 0.9% physical violence, and 0.9% sexual violence. This highlights the need for policies to prevent workplace violence, as recommended by 92.9% of participants.(26)

### Implications for Medical Education and Policy

Given how rampant the issues are and how much they affect the lives of HCWS, the role of medical faculty in fostering discussions and debates, creating awareness, and educating one another cannot be overemphasised. It is obligatory that they identify, prevent, and address the factors contributing to this rampant spike in violence. At the institution where the study was conducted, student researchers were encouraged to engage in discussions during class about the State Act to educate and speak about the Act during class sessions, allowing them to educate their peers. However, the impact would have been wider if students had dedicated time and the institutional support for structured awareness sessions, enabling greater participation, from faculty and senior staff. This underscores the crucial societal role of educational institutions in encouraging engagement with real-world events, as medical professionals cannot remain isolated from societal and systemic challenges.

During the awareness class, when asked about a recent violent experience they had witnessed, heard about, or gone through, most of participants cited the physical assault on a medical resident that took place at Gandhi Hospital in Telangana.(27) Additionally, some participants recalled other instances of attacks on doctors, in cities such as Vijayawada(28), Kolkata(29), Indore(30), etc. None of the students cited occurrences involving verbal abuse, damage to clinical facilities, or other forms of violence; all responses discussed cases of physical assault against doctors.

According to 87.9% of participants, existing laws do not adequately protect doctors from assault, despite the Kerala Act’s provisions. This aligns with findings by Gohil RK et al (2018), where only 20% of violence cases were reported to the police and 56.3% of offenders faced no consequences whereas 23.9% were issued a verbal warning. (31) The lack of enforcement suggests that legal protections must not only be enforced but their provisions, widely disseminated. The majority of participants, 85.2%, believed that all forms of violence against doctors, including psychological abuse, should be thoroughly examined, condoned, and dealt with appropriately through law enforcement, and through the judiciary.

Medical educators have the responsibility to take a proactive role in educating themselves about medical laws and creating awareness among students. Through this research, which was a part of their curriculum, students engaged in discussions about the Act during class, but institutional barriers such as limited time and space for awareness sessions to faculty through Clinical Clubs, restricted broader participation. Strengthening faculty-led discussions, debates, and practical case-based learning with multisectoral participation, could further institutionalize legal education and medical jurisprudence. Ranjan et al., quoted that a majority of students gathered knowledge about increasing violence against HCWs through media (91.6%) rather than formal education, thereby revealing a pressing need for structured legal education through medical curricula.(25)

### Bridging the gap between Awareness and Practice

Collective awareness and engagement can bring meaningful changes to safeguard medical professionals. Discussions on the protection of healthcare workers, particularly issues affecting the safety of their community, should not be silenced, or dismissed. The notion that such concerns are irrelevant unless personally affected fosters a culture of complacency. Instead, open discourse should be encouraged at all levels, without fear or hesitation, to ensure that these critical issues receive the attention they deserve.

Legal provisions such as the “Kerala Healthcare Service Persons And Healthcare Service Institutions (Prevention Of Violence And Damage To Property) Amendment Act, 2023” exist but systemic barriers prevent the effective enforcement of such laws. This study reinforces the fact that curricular educational interventions improve knowledge, but for real-world application, a multi-pronged approach is necessary. Even success of the “Code Grey” protocol for Kerala hospitals, though a laudable initiative, depends on consistent training and HCW participation.(32)

### Global Perspectives on Healthcare Worker Protection Laws

Comparing Kerala’s Act with international frameworks highlights the best framework within which such laws could work. The Workplace Violence Prevention for Health Care and Social Service Workers Act (2021) focuses on proactive preventive action rather than reflex, punitive action after the event, in the United States. (33)In the United Kingdom, Assaults on Emergency Workers (Offences) Act of 2018, has successfully improved legal awareness. In China, another country affected by healthcare workers violence, limited awareness of legal recourse hinders the impact. (34). The Kerala Protection Act though a step forward is more punitive than preventative, highlighting the need for integrated training programs, an amalgamation of the interface between medicine and law, and systemic reforms to strengthen HCW protections.

### Limitations

While this study demonstrated a significant improvement in awareness and knowledge of the Act post-intervention, its generalizability remains a concern. Conducted at a single medical college, the findings may not reflect students from other regions with different curricula and exposure to healthcare policies. Additionally, the study focused only on medical students, excluding other healthcare professionals whose awareness levels and learning needs may differ. Knowledge retention from a single intervention is also uncertain without reinforcement.

### Recommendations for Interventions

The study recommends integrating structured awareness programs on healthcare-related legislation into medical curricula to ensure sustained knowledge acquisition among students. Regular follow-up sessions, workshops, and refresher courses could reinforce this knowledge and promote long-term retention. Mandatory legal training sessions in collaboration with legal experts and law enforcement as part of their initiation programme to ensure that all Healthcare Workers are aware of this Act and their rights. Pamphlets, posters, and digital works should be made available in the health care setting. Association with legal bodies to conduct workshops which would allow them to address their concerns. Similar interventions should also target other healthcare professionals, including nurses, paramedics, and emergency staff, to address the needs of those actively facing workplace violence. Simulation-based training can further enhance the application of legal knowledge by providing students and professionals with practical experience in handling real-world scenarios.

### Future Research

Future research should focus on longitudinal studies to evaluate the long-term retention of knowledge and its impact on behaviour and decision-making in clinical practice. Multi-centre studies involving participants from various regions and institutions are necessary to enhance the generalizability of findings. Another area of investigation can explore the link between increased awareness of the Act and measurable behavioural changes, such as improved incident reporting or readiness to handle workplace violence. Comparative studies on different educational approaches, such as in-person classes, e-learning modules, and blended learning, can help identify the most effective methods for delivering legal education. Additionally, research should assess the awareness and attitudes of high-risk groups, such as emergency department staff, and evaluate how workplace culture and administrative support influence the practical application of legal knowledge. Further studies could also examine the role of community and patient education in reducing violence against healthcare workers and investigate how organizational factors, such as reporting systems and security measures, impact the success of such interventions. Together, these research directions can provide valuable insights into creating more effective strategies for mitigating workplace violence in healthcare settings.

In conclusion, while this study has promising results in improving knowledge through targeted educational interventions, a broader institutional commitment to legal education, and policy enforcement is mandated. Further research to assess the broader applicability of these findings and the long-term impact of such interventions in real-world settings is needed. While Kerala’s legislative effort is commendable, its impact among healthcare professionals across various healthcare institutions in Kerala—and perhaps expanding this to other regions— will remain limited, unless awareness, training, and enforcement mechanisms are systematically strengthened. By bridging the gap between knowledge and action through sustained education, inclusive training, and systemic support, can healthcare workers be empowered to navigate challenges confidently and foster safer, violence-free healthcare environments. Ultimately, societal change that promotes respect, understanding, and empathy toward healthcare workers will go a long way to reducing violence and ensuring a safer, supportive environment for both healthcare providers and patients.

## Data Availability

All data produced in the present study are available upon reasonable request to the authors.

